# Saliva as a Reliable Sample for COVID-19 Diagnosis in Paediatric Patients

**DOI:** 10.1101/2021.03.29.21254566

**Authors:** Alvina C. Felix, Anderson V. de Paula, Andreia C. Ribeiro, Francini C. da Silva, Marta Inemami, Angela A. Costa, Cibele O. D. Leal, Walter M. Figueiredo, Dmitry J. S. Sarmento, Tatiana A. Sassaki, Claudio S. Pannuti, Paulo H. Braz-Silva, Camila Malta Romano

## Abstract

Saliva has been described a less invasive and easy to handle sample, compared to nasopharyngeal swabs (NPS), in the diagnosis of COVID-19 in adults. Although the advantages of using saliva is still more evident in paediatric patients, little is now about its sensitivity in this group. The aim of this study was to compare the performance of saliva to that of NPS in the detection of SARS-CoV-2 in paediatric patients with mild symptoms. This study evaluated saliva samples from children with suspected COVID-19 who attended public healthcare services of Araraquara, São Paulo, Brazil. Children were asked to spit into a sterile container for collection of about 1ml of saliva after the NPS collection. SARS-COV-2 detection was performed by using the Altona RealStar® SARS-CoV-2 RT-PCR Kit 1.0. The sample consisted of 50 patients, in which 27 were girls (54%) and 23 were boys (46%). Ten were positive for SARS-CoV-2 in at least one sample collected. The mean age was 10.24 ± 3.52 years old and saliva was collected after 4.76 ± 1.31 days from the symptoms. Saliva and NPS have showed the same performance in the SARS-CoV-2 detection (k = 0.865, P < 0.001). In conclusion, saliva is a reliable alternative sample for COVID-19 diagnosis in paediatric population.

## Introduction

Massive testing is one of the most effective strategies for preventing COVID-19 transmission by allowing early identification of cases and decision-making based on the pandemic’s behaviour^1^. Nasopharyngeal swab (NPS) is the standard sample for molecular test, which causes much discomfort and possible cross-contamination during the collection^1,2^. Therefore, a trained healthcare professional is required for performing this procedure^1,3^. Saliva has been shown to be a reliable, safe diagnostic fluid for detection of SARS-CoV-2, with a sensitivity ranging from 80-100% compared to NPS in adult population^1,3,4^. In paediatrics, the use of saliva has clear advantages because the collection of this fluid is less invasive, thus reducing the discomfort and allowing self-collection^2^.

The objective of the present study was to compare the performance of saliva to that of NPS in the detection of SARS-CoV-2 in paediatric patients with mild symptoms.

## Material and Methods

This study evaluated saliva samples from children with suspected COVID-19 who attended public healthcare services of Araraquara, which is a medium-sized city located in the State of São Paulo, with a population of 238,339 in 2020.

As part of the COVID-19 contingency plan, the city of Araraquara offers molecular tests for detection of SARS-COV-2 by using NPS in all symptomatic patients seeking healthcare service.

Parents and their children were invited to participate in this study at the time of NPS collection, in which the children were asked to spit into a sterile container for collection of about 1ml of saliva. The saliva samples were immediately stored at 4°C until being taken to the laboratory (< 48 hours). Symptoms and delay between their onset and sample collection (days) were also recorded. Total RNA was extracted by using the viral RNA mini kit (Qiagen, GE) and SARS-COV-2 detection was made by using the Altona RealStar® SARS-CoV-2 RT-PCR Kit 1.0 (Altona Diagnostics GmbH, Hamburg, Germany). The positivity in saliva was later compared to the results of NPS obtained by the Araraquara health surveillance. Unfortunately, no information on viral load or threshold cycle (CT) in the NPS test was available for further comparisons.

This study was approved by the Research Ethics Committee of the University of São Paulo School of Medicine under protocol number 4235245.

## Results

The sample consisted of 50 patients, in which 27 were girls (54%) and 23 were boys (46%). Ten were positive for SARS-CoV-2 in at least one sample collected (saliva or NPS). The mean age was 10.24 ± 3.52 years old and saliva was collected after 4.76 ±

1.31 days from the symptoms. Of the 50 patients evaluated, symptoms were reported by 46 during the saliva collection and the main ones were the following: coryza (60.9%), cough (56.5%), sore throat (45.7%), headache (39.1%) and fever (30.4%). None of these symptoms was statistically associated with the diagnosis of COVID-19 (Table 1).

**Table 1.** List of Symptoms

| Symptoms |  | n | % | *p*(1) |
| --- | --- | --- | --- | --- |
| Fever | Yes | 14 | 30.4 | 0.242 |
|  | No | 32 | 69.6 |  |
| Cough | Yes | 26 | 56.5 | 0.150 |
|  | No | 20 | 43.5 |  |
| Shortness of air | Yes | 9 | 19.6 | 0.384 |
|  | No | 37 | 80.4 |  |
| Coryza | Yes | 28 | 60.9 | 0.717 |
|  | No | 18 | 39.1 |  |
| Headache | Yes | 18 | 39.1 | 0.999 |
|  | No | 28 | 60.9 |  |
| Myalgia | Yes | 7 | 15.2 | 0.163 |
|  | No | 39 | 84.8 |  |
| Fatigue | Yes | 8 | 17.4 | 0.664 |
|  | No | 38 | 82.6 |  |
| Nausea | Yes | 4 | 8.7 | 0.201 |
|  | No | 42 | 91.3 |  |
| Vomit | Yes | 4 | 8.7 | 0.999 |
|  | No | 42 | 91.3 |  |
| Diarrhea | Yes | 6 | 13.0 | 0.315 |
|  | No | 40 | 87.0 |  |
| Abdominal pain | Yes | 8 | 17.4 | 0.055 |
|  | No | 36 | 82.6 |  |
| Loss of the sense of smell | Yes | 0 | 0 | - |
|  | No | 46 | 100 |  |
| Loss of the sense of taste | Yes | 2 | 4.3 | 0.391 |
|  | No | 44 | 95.7 |  |
| Sore throat | Yes | 21 | 45.7 | 0.306 |
|  | No | 25 | 54.3 |  |

<sup>(1)</sup> Association of the symptom with diagnosis of COVID-19. Fisher's exact test.

ROC curve analysis was performed in order to assess sensitivity and specificity of RT-PCR between saliva and NPS, with the latter being considered a gold standard test. The results showed a statistically significant curve (AUC = 0.851, SE = 0.094; *P* = 0.002; 95% CI = 0.667 – 1.00). With these results, we can state that saliva can be safely used for diagnosis of COVID-19 in paediatric patients. We also tested the concordance between saliva and NPS by using Kappa concordance test (k = 0.702; *P* < 0.001), with 92% of the samples being concordant (Table 2). Additionally, the concordance between these fluids was assessed individually, in which positive cases (10/50 patients) were considered as true infection. It was observed that the saliva and NPS showed the same values for Kappa concordance test (k = 0.865, *P* < 0.001).

**Table 2.** Description of positive and negative cases of COVID-19 for saliva and NPS by using RT-PCR.

|  |  | NPS |  | Total |
| --- | --- | --- | --- | --- |
| RT-PCR |  | Positive | Negative |  |
|  |  | N(%) | N(%) |  |
| Saliva | Positive | 6 (12) | 2 (4) | 8 (12) |
|  | Negative | 2 (4) | 40 (80) | 42 (84) |
| Total |  | 8 (12) | 42 (84) | 50 (100) |

## Discussion

Consistent scientific evidence has pointed to the effectiveness of the use of saliva as a diagnostic fluid for COVID-19 in adult population^1, 3, 4^. The advantages of a less invasive and painless sample collection have been shown to be more evident in paediatric population, which may include self-collection and multiple collection possibilities^2^. However, there are a few studies of paediatric patients and different ways to sampling for SARS-CoV-2 detection and their results are conflicting^5-7^. Our results showed that saliva had the same diagnostic performance than that of NPS for SARS-CoV-2.

The use of less invasive strategies for COVID-19 surveillance has a crucial importance for children not only in the understanding of SARS-CoV-2 behavior in this population, but also in the re-opening of schools based on constant tracking of asymptomatic cases^2,6^. Additionally, with the emergence of the variants of concern and vaccination campaigns were initially not aimed at this age group, saliva could help on the detection of them in paediatric patients. The use of saliva makes COVID-19 surveillance viable, since this strategy is more largely accepted by individuals for being painless and for requiring no professional sample collection^2,3^. Depending on the age group, the individuals themselves can be remotely instructed (e.g. videos) to perform the sample collection^3^.

The limitations of this preliminary work are in the fact that larger samples and inclusion of asymptomatic children should be considered in further studies.

## Conclusion

Our data allow us to conclude that saliva is a viable alternative fluid for molecular diagnosis of COVID-19 in children.

## Data Availability

Data availability under request.

## Acknowledgments

The authors would like to thank José Tadeu Sales for the language correction of the manuscript. This study was supported by the São Paulo Research Foundation (FAPESP) according to grant # 2019/03859-9

## References

1. Sapkota D, Søland TM, Galtung HK, Sand LP, Giannecchini S, To KKW, et al. COVID-19 salivary signature: diagnostic and research opportunities. J Clin Pathol 2020;Aug7:jclinpath 2020–206834. doi: 10.1136/jclinpath-2020-206834.

2. Santos CN, Rezende KM, Oliveira Neto NF, Okay TS, Braz-Silva PH, Bönecker M. Saliva: an important alternative for screening and monitoring of COVID-19 in children. Braz Oral Res 2020;34:e0125. doi: 10.1590/1807-3107bor-2020.vol34.0125.

3. Braz-Silva PH, Mamana AC, Romano CM, Felix AC, de Paula AV, Fereira NE, et al. Performance of at-home self-collected saliva and nasal-oropharyngeal swabs in the surveillance of COVID-19. J Oral Microbiol 2020;13(1):1858002. doi:10.1080/20002297.2020.1858002.

4. To KK, Tsang OT, Yip CC, Chan KH, Wu TC, Chan JM, et al. Consistent Detection of 2019 Novel Coronavirus in Saliva. Clin Infect Dis 2020;71(15):841–843. doi: 10.1093/cid/ciaa149.

5. Kam KQ, Yung CF, Maiwald M, Chong CY, Soong HY, Loo LH, et al. Clinical Utility of Buccal Swabs for Severe Acute Respiratory Syndrome Coronavirus 2 Detection in Coronavirus Disease 2019-Infected Children. J Pediatric Infect Dis Soc 2020;9(3):370–372. doi:10.1093/jpids/piaa068.

6. Chua GT, Wong JSC, To KKW, Lam ICS, Yau FYS, Chan WH, et al. Saliva viral load better correlates with clinical and immunological profiles in children with coronavirus disease 2019. Emerg Microbes Infect 2021;10(1):235–241. doi: 10.1080/22221751.2021.1878937.

7. Chan RWY, Chan KC, Chan KYY, Lui GCY, Tsun JGS, Wong RYK, et al. SARS-CoV-2 detection by nasal strips: A superior tool for surveillance of paediatric population. J Infect 2020;S0163-4453(20)30704-0. doi: 10.1016/j.jinf.2020.11.009. Epub ahead of print.

